# Quantifying risk modifiers of hereditary hemochromatosis using genomic and electronic health record data from FinnGen and UK Biobank

**DOI:** 10.1101/2025.09.30.25336973

**Authors:** Jarkko Toivonen, Jonna Clancy, FinnGen, Fredrik Åberg, Jarmo Ritari, Mikko Arvas

## Abstract

Hereditary hemochromatosis is an autosomal recessive disorder characterized by excessive iron accumulation in the body. Early diagnosis of hemochromatosis allows starting treatment before severe organ damage has occurred. The C282Y variant in the *HFE* gene is the most common cause of hemochromatosis. However, its penetrance of only 20% limits its utility for population-wide screening. We aimed to identify and quantify the predictive value of novel genetic variants and non-genetic factors from electronic health care records that could modify the effect of the C282Y variant on hemochromatosis, thereby partly explaining its incomplete penetrance.

We carried out a cohort study on data from 420,543 individuals in the FinnGen project, for whom genotype information and health care records were available. We performed a regular and an interaction GWAS of hemochromatosis and fitted statistical models of hemochromatosis with multiple genetic and non-genetic predictors. We validated our results using data from the UKBB.

Our analyses identified one novel statistically significant variant (rs181949568) in *CASC15* gene, within 4 Mb from the *HFE* gene, and we were able to quantify the strong protective role of blood donation.

We found that donating at least twice a year is likely to be sufficient to drop the risk of C282Y homozygote to that of a C282Y-H63D compound heterozygote. To facilitate clinical application, we present the results as individual-level risk tables. Also, we found that in Finland hemochromatosis related diagnosis predicted later hemochromatosis diagnosis suggesting that in Finland hemochromatosis is not well recognized.

## Introduction

Hereditary hemochromatosis is an autosomal recessive disorder in which excess iron accumulates in organs such as liver, heart and joints. As iron in certain forms can cause oxidative stress, this buildup can lead to organ damage. Common consequences of hemochromatosis include liver disease, liver cirrhosis, congestive heart failure, diabetes, and arthritis (1). The prevalence of hemochromatosis is highest in populations of Nordic or Celtic origin, with approximately one in every 220 to 250 individuals diagnosed (2).

The C282Y variant of the *HFE* gene is assumed to downregulate the activity of hepcidin, whose absence in turn allows uninhibited release of iron from enterocytes and macrophages into circulation (3). Even though the homozygous C282Y variant has a prevalence of about 5 in 1000 individuals of Northern European descent (4), only a small fraction of these individuals develops the disease. For instance, in the UK Biobank (UKBB) it was estimated that the penetrance in C282Y homozygotes was 25.3% for males and 12.5% for females (5). On the other hand, by following incidences up to age of 80, the cumulative incidences of 56.4% and 40.5% for males and females, respectively, have been reported (6).

It is important to diagnose the disease early, so that treatment can be started before significant organ damage has occurred. Common and effective treatment is removal of excess iron through venesections. Blood donation can also be used as treatment in the maintenance phase in case no organ damage has occurred. Blood donation could also prevent organ damage among C282Y homozygotes, as in a recent study no indication of organ damage was detected in frequent blood donors homozygous for C282Y (FRCBS, unpublished data).

Previously, Gallego et al have compared the penetrance of C282Y homozygotes and C282Y-H63D compound heterozygotes in the American eMERGE cohort (7) and found signicant difference in diagnosis rates and among males in clinical penetrance. Pilling et al have investigated the effect of *HFE* variants on the baseline prevalence and incidence of hemochromatosis related diseases, and found significant difference in incidence of associated conditions during 7-year follow-up and among males in prevalence of associated conditions, when comparing C282Y homozygotes and C282Y wild types(8). They have also studied the effect of genetic scores of four iron markers, liver cirrhosis, osteoarthritis and type 2 diabetes on the penetrance of *HFE* variants on hemochromatosis related diseases and found significant associations between the scores and related diseases among C282Y homozygotes (5).

In this paper we use genetic and health care register data to model the incidence and severity of hereditary hemochromatosis in the FinnGen cohort of 500,000 Finnish individuals. Our aim is to discover new genetic variants and their interactions that could explain the low penetration of the *HFE* mutations, and thereby to assess whether genetic screening for hemochromatosis risk SNPs is warranted in Finland. We discover new SNPs and interactions having effect on the diagnosis of hemochromatosis and present our findings as tables characterizing the personal-level hemochromatosis risk.

## Methods

### Materials

The FinnGen project (9) recruited 520,210 individuals, of age at least 18, within period of 2017–2023 through various Finnish biobanks. For each participant 21,331,644 imputed genetic variants, various laboratory measurements and other health register data are available.

The UK biobank (UKBB) (10) recruited circa 500,000 individuals, aged 40-69, between years 2006– 2010. For each participant genomic data, consisting of 96 million variants, large amount of questionnaire, omics, and health care register data are available.

In FinnGen the hemochromatosis diagnosis (endpoint E4_IRON_MET) is based on the ICD-10 code E83.1 and the ICD-9 code 2750. Individuals with metabolic disorders (ICD-10 codes E70–E90) are excluded from controls. The diagnoses come from the death register (follow-up from 1969 to 2021) and hospital discharge register (follow-up for inpatient and outpatient registers are 1969–28.4.2023 and 1998–28.4.2023, respectively). The diagnosis of primary *HFE* hemochromatosis in Finland is given to a patient with transferrin saturation above 45%, ferritin above the lab- and sex-specific reference limit and *HFE* genotype either C282Y homozygote or C282Y/H63D heterozygote (11). In the UKBB data we define hemochromatosis similarly based on the ICD-10 codes.

Since menstruation protects from hemochromatosis due to blood loss, it is important to include the menopausal status in the hemochromatosis models. We define females at most 50 years old as premenopausal.

We use the number of venesections performed as the measure of severity of hemochromatosis. The Nomesco code TPH00 and the SPAT code (Finnish primary care outpatient procedures) SPAT1074 give the venesection procedures in FinnGen. However, we excluded individuals with either polycythemia vera (1236 individuals) or porphyria cutanea tarda (35 individuals) diagnosis from the disease severity analyses as venesection is a treatment for these diseases as well.

Blood donation histories for 48,243 individuals are comprehensively available from years 2000 to 2021. We use the number of donations in the previous five years as a predictor of hemochromatosis incidence, survival and disease severity.

The FinnGen cohort contains 520,210 individuals, and after exclusions, described in Figure 1, 420,543 individuals remain. The UKBB includes 502,547 individuals, and after exclusions 318,778 individuals remain. Individuals with missing information were filtered out.

**Figure 1.**
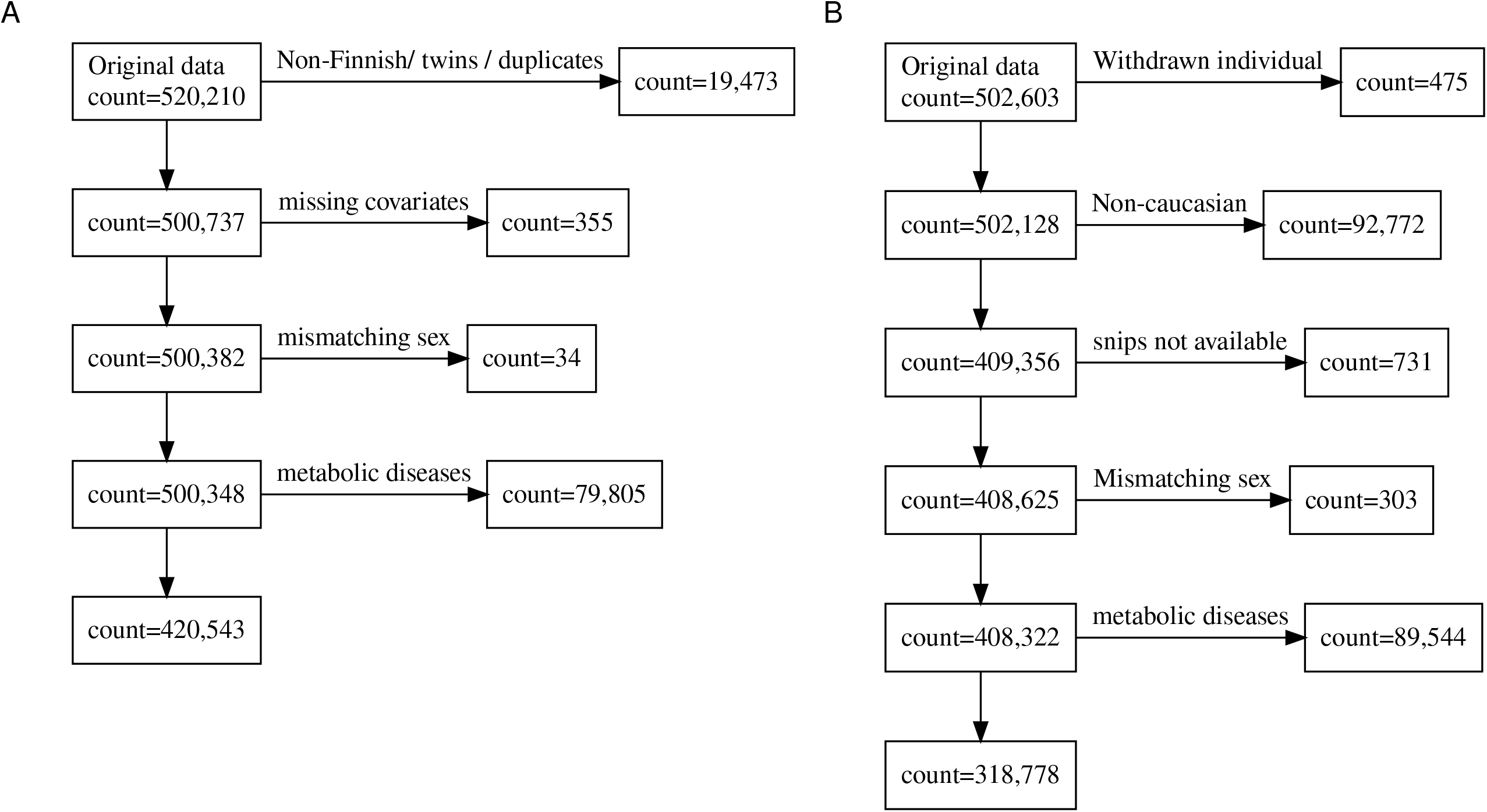
The exclusions performed in A) the FinnGen cohort and in B) the UKBB cohort.

### Statistical analyses

The additive hemochromatosis GWAS is done using Regenie, using age, sex, genotyping batch and the first 10 principal components as covariates on FinnGen data. Fine-mapping of causal variants is done using Susie. The extended MHC region (GRCh38, chromosome 6, 25–34 Mb) was excluded from fine-mapping due to the complex LD structure of the area. The association study between hemochromatosis and the alleles of 15 HLA genes and the genome-wide search of putative variants that additively interact with C282Y were performed with Regenie.

We model the individual-level hemochromatosis risk as the probability of hemochromatosis diagnosis at the end of follow-up given by a binary response logistic regression or XGBoost classification model, with the latter having the benefit of automatically selecting important variables and their interactions as well as the functional form of the dependence of the response. The survival from hemochromatosis is modeled using Cox proportional hazards model. We use the number of venesections performed as a proxy for disease severity, although the venesections from the maintenance phase may make this estimate of disease severity slightly biased. The venesection count response is modeled using either Poisson or negative binomial model; or their zero-inflated versions, since most individuals have had no venesection treatments, and even of hemochromatosis cases less than half have received venesection.

We specify three versions of the incidence and survival models, differing by the set of predictors used: a simple baseline model with only C282Y and H63D as genetic variables; a comparison model that consists of all relevant genetic and non-genetic prediction variables in common with FinnGen and UKBB; and the best FinnGen model that contains all relevant genetic and non-genetic FinnGen predictors. We included interactions of C282Y with all other predictors that do not induce collinearity among the predictors (variance inflation factor < 5). All the SNPs are modeled additively, except for C282Y and H63D, which were modeled as categorical variables, due to them showing non-additive effects in previous literature. This allows all nine combinations of these two variants. In the best FinnGen model we also include correlated previous diagnoses of other diseases as predictors, see the Supplement on the selection of the used diseases. For the disease severity model, we use the same predictors as in the Best FinnGen model except the preceding diagnoses were excluded to avoid fitting problems since the venesections are quite rare.

Best models are selected based on performance on held-out data, see Supplemental methods. A fixed-effect meta-analysis is performed between effect sizes of comparison models of FinnGen and UKBB.

As a sensitivity analysis we refitted the Comparison model and the Best FinnGen model in data where cases were required to have at least one venesection procedure in addition to the register diagnosis of hemochromatosis.

Further details, including the used R packages and their versions, can be found in the Supplemental methods.

## Results

The demographic information about the FinnGen and UKBB cohorts after exclusions are shown in Table 1. The hemochromatosis prevalence in UKBB is about 2.4 times that of FinnGen. Also, the mean age of participants at baseline was higher in UKBB, due to the minimum recruitment age of 40. On the other hand, the male-female ratio was more even in the UKBB, while clear majority of the participants were female in both cohorts.

**Table 1.**
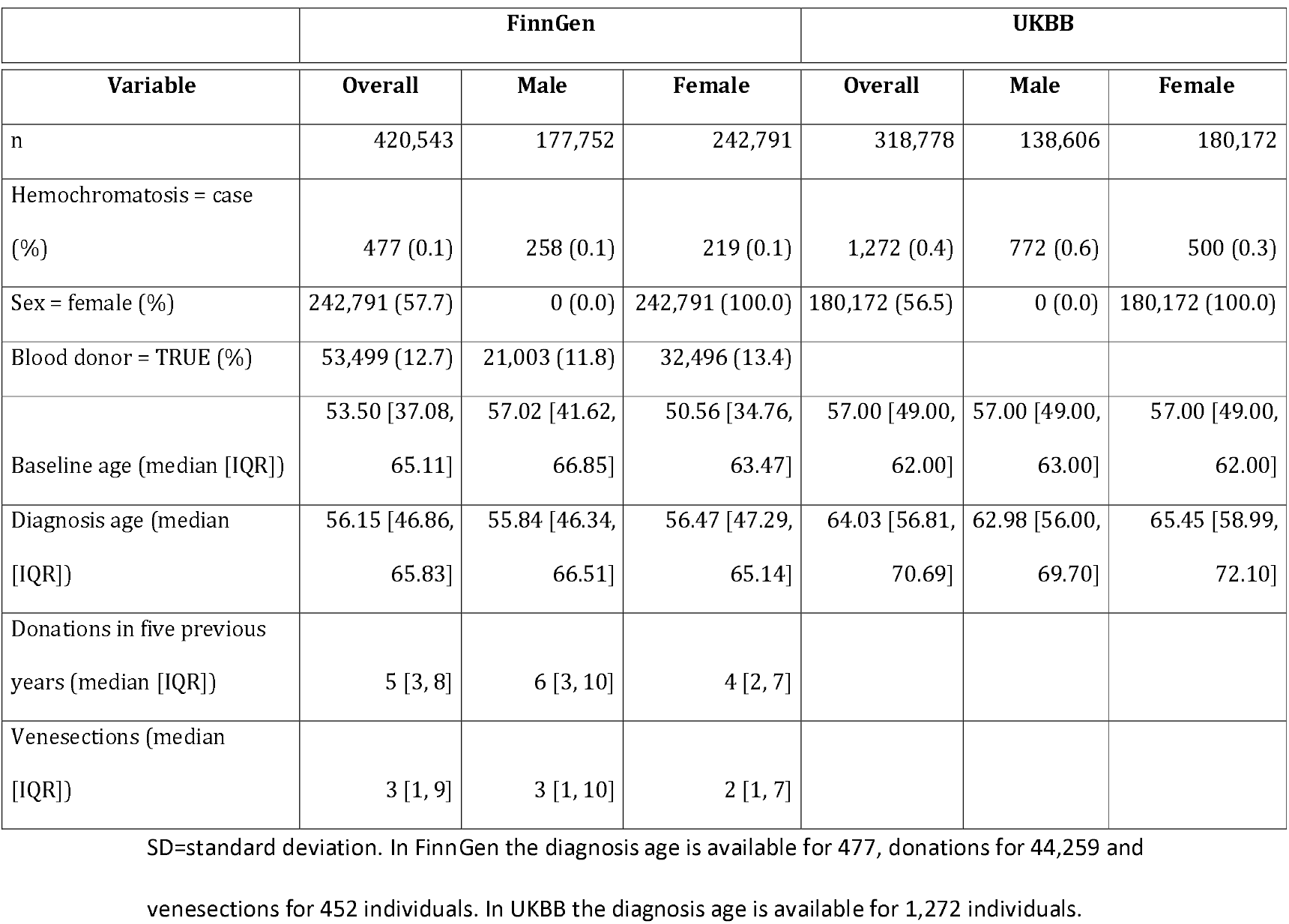
Demographics statistics (after exclusions have been performed) in FinnGen and UKBB cohorts.

| Variable | Overall | Male | Female | Overall | Male | Female |
| --- | --- | --- | --- | --- | --- | --- |
| n | 420,543 | 177,752 | 242,791 | 318,778 | 138,606 | 180,172 |
| Hemochromatosis = case (%) | 477 (0.1) | 258 (0.1) | 219 (0.1) | 1,272 (0.4) | 772 (0.6) | 500 (0.3) |
| Sex = female (%) | 242,791 (57.7) | 0 (0.0) | 242,791 (100.0) | 180,172 (56.5) | 0 (0.0) | 180,172 (100.0) |
| Blood donor = TRUE (%) | 53,499 (12.7) | 21,003 (11.8) | 32,496 (13.4) |  |  |  |
| Baseline age (median [IQR]) | 53.50 [37.08, 65.11] | 57.02 [41.62, 66.85] | 50.56 [34.76, 63.47] | 57.00 [49.00, 62.00] | 57.00 [49.00, 63.00] | 57.00 [49.00, 62.00] |
| Diagnosis age (median [IQR]) | 56.15 [46.86, 65.83] | 55.84 [46.34, 66.51] | 56.47 [47.29, 65.14] | 64.03 [56.81, 70.69] | 62.98 [56.00, 69.70] | 65.45 [58.99, 72.10] |
| Donations in five previous years (median [IQR]) | 5 [3, 8] | 6 [3, 10] | 4 [2, 7] |  |  |  |
| Venesections (median [IQR]) | 3 [1, 9] | 3 [1, 10] | 2 [1, 7] |  |  |  |

SD=standard deviation. In FinnGen the diagnosis age is available for 477, donations for 44,259 and venesections for 452 individuals. In UKBB the diagnosis age is available for 1,272 individuals.

### GWASes

A Manhattan plot of hemochromatosis GWAS on FinnGen data (477 cases and 420,066 controls) is shown in Figure S1, QQ-plot in Figure S2 and lead variants in Table S1. The Manhattan plot included only one peak, in chromosome 6 at the *HFE* gene, ranging approximately from 21.9 Mb to 34.1 Mb. Fine-mapping of the GWAS hits produced three high-quality credible sets, whose representative SNPs are listed in Tables S2. These representative SNPs were chosen for further statistical analyses (SNPs Causal1 to Causal3).

As the extended HLA region was excluded from fine-mapping, we also selected all independent significant lead SNPs from the HLA area. This resulted in SNPs HLA1 and HLA2 for further analysis, see Supplemental results and Figure S3. Finally, we selected SNPs C282Y, H63D (12), S65C (13), H1-2 and HLA3 (14) based on previous literature.

As the HLA genes were not associated with hemochromatosis when adjusting with C282Y, we did not include any HLA alleles in the statistical models, see Supplemental results for details.

We also used Regenie to find putative interactions with C282Y that could be associated with hemochromatosis. This GWAS performed was on the chromosome 6 only, since all the significant associations in the basic GWAS appear in that chromosome, see Figure S5.A. All the significant interactions with C282Y were found in region 25,897,390–26,135,271 Mb. The significant interactions included the known interactions C282Y-H63D and C282Y-H1-2. Since we wanted to find new interactions, we performed the GWAS again, this time adjusted with the SNPs H63D, S65C, H1-2 and HLA3, and restricted to range chr6:24.8–29.7 Mb (see Figure S5.B and Table S3), resulting in 67 significant SNPs. We tested the utility of these SNPs in a later statistical model (the All interactions model) of hemochromatosis incidence.

### SNPs

The selected SNPs, except the 67 C282Y-interaction SNPs, are described in more detail in Table S4 and Figure 2, stratified by the hemochromatosis status.

**Figure 2.**
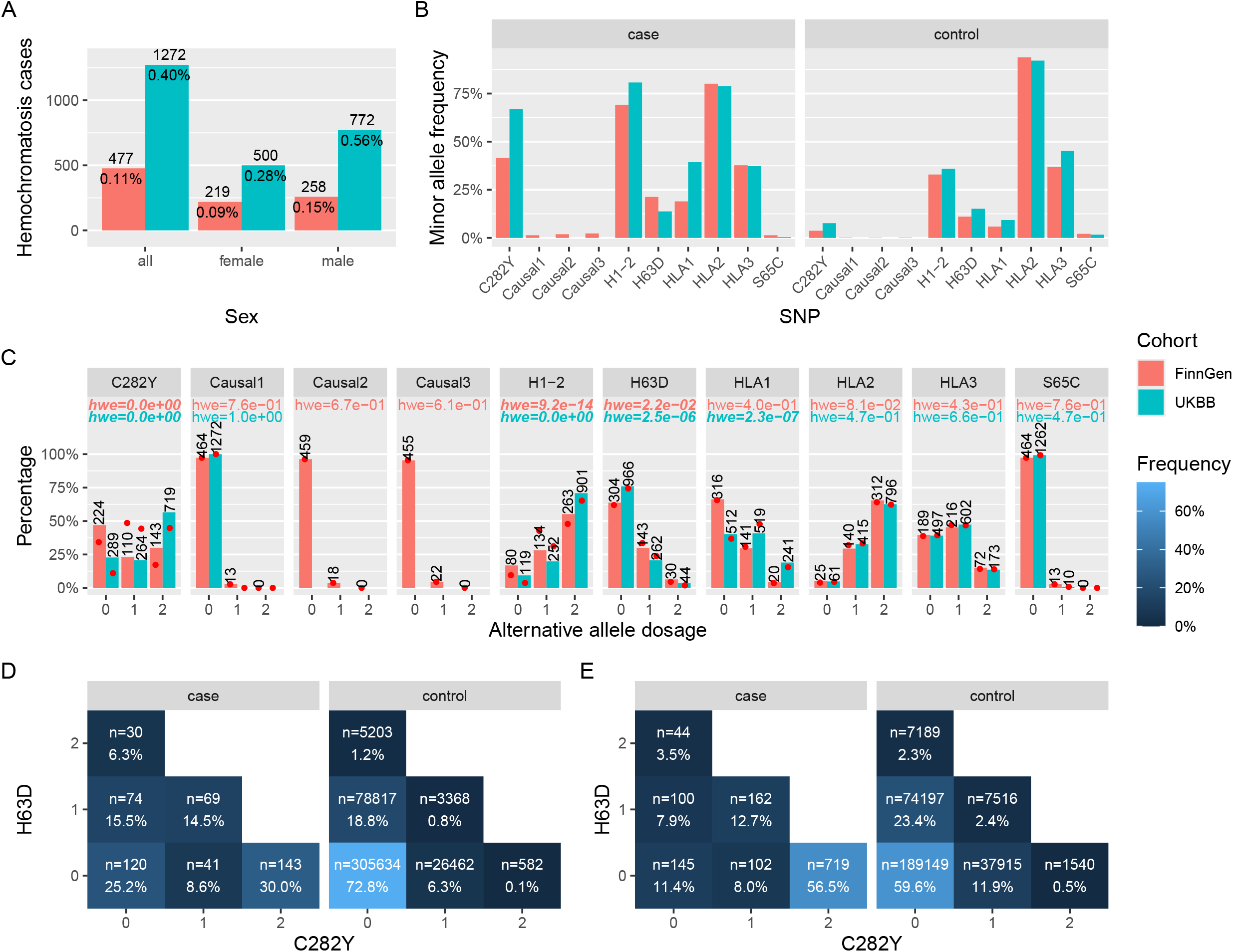
Information on the genetic variants putatively related to a registry diagnosis of hemochromatosis. A) Hemochromatosis prevalence in FinnGen and UKBB stratified by sex. B) Minor allele frequencies stratified by the hemochromatosis status. C) Observed minor allele dosages as bars, dosages expected based on MAF in red dots and the HWE test result among cases. The p-values of SNPs significantly deviated from equilibrium are indicated in bold and italic. D) Dosage combinations of C282Y and H63D occurring in FinnGen. E) Dosage combinations in UKBB.

As can be seen in Figure 2.A the HC prevalence was over twice in UKBB than that in FinnGen. Also, hemochromatosis was more common among males, in both FinnGen and in UKBB. In the minor allele frequencies (MAFs) there were larger differences between cases and controls, especially in C282Y and H1-2, except for H63D in the UKBB and HLA3 in FinnGen. The variants C282Y, H1-2, and H63D are not in HW equilibrium among cases, possibly indicating their importance in predicting the future hemochromatosis diagnosis. From panels D and E, we can also see that when an individual is homozygote for C282Y, then the individual does not carry an H63D mutation, and vice versa. So, likely the C282Y and H63D alleles cannot be in the same chromosome.

In FinnGen 75% of hemochromatosis cases carry at least one mutation in C282Y or H63D, while in UKBB the fraction is 88.6%. On the other hand, the penetrance of the homozygote C282Y and the compound heterozygote C282Y-H63D in FinnGen is 20.2% and 2%, respectively, while in UKBB the corresponding numbers are 24.8% and 1.6%.

### Performances and model selection

As the more complicated models of hemochromatosis incidence did not significantly improve performance (see supplemental results), we concentrated on the logistic regression based models for further analysis. Poisson model, negative binomial model, and their zero-inflated versions were experimented with for the disease severity, and zero-inflated Poisson model was selected, based on the AIC values and the instability of the other models on the data.

### Inference

The odds ratios of the meta-analysed logistic comparison model and the best FinnGen model; the hazards ratios of the meta-analysed survival comparison model and the best FinnGen model; and the incidence ratios of the venesection model are visualized in Figure 3 (also in Table S6), along with the corresponding 95% confidence intervals. The variable importances are shown in Figures S8 and S9, and in Tables S7 and S8.

**Figure 3.**
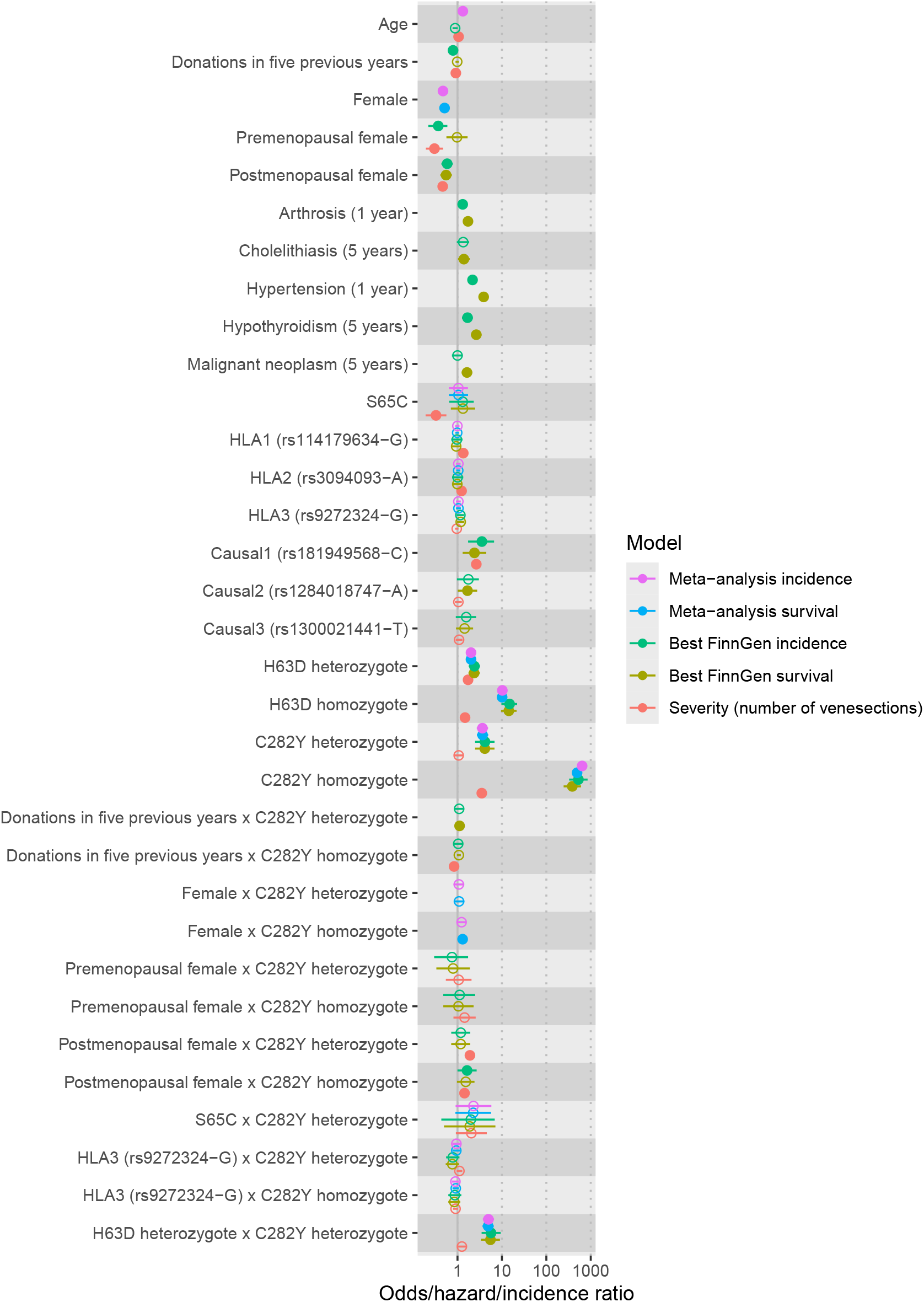
Effect sizes of the hemochromatosis incidence (odds ratio from logistic regression), survival (hazard ratio from Cox model) and severity models (incidence ratio). Full data was used to fit the models. For disease severity the number of venesections were modeled using zero-inflated Poisson model with same predictors for both the count and zero components. The 95% confidence intervals are shown as horizontal lines. The minimum gap in years to previous disease diagnoses is indicated in parentheses. The effect alleles for the novel variants are given after the RSIDs. The age at the end of follow-up was standardized with unit given in standard deviations.

Both logistic and Cox versions of the best FinnGen model seem to have similar effect sizes, except for the preceding diagnoses, the number of blood donations in the last five years and being premenopausal female. Also, the meta-analyses of the logistic comparison models and Cox comparison models seem to produce similar effect sizes. The venesection model gave considerably differing effect size estimates compared to the rest of the models for predictors Age, C282Y heterozygote and homozygote, compound heterozygote of C282Y and H63D, H63D homozygote and S65C.

Although the effect size of Donations in five years is low, note that the corresponding unit is one donation. Hence, if a male has donated the maximum of 30 times in five years, the odds ratio would be raised to the power of 30. For example, for the odds ratio of 0.9 for a single donation the odds ratio for 30 donations in five years would be 0.9^30^ ≈ 0.04, which is the biggest protective effect over all predictors.

All the diagnoses of malignant neoplasm, hypothyroidism, hypertension, cholelithiasis and arthrosis preceding the first hemochromatosis event (see Supplemental Table S9 for the required minimum time window between the disease and hemochromatosis event) were risk inducing. Note that the venesection model has less predictors than the best FinnGen model since the small number of venesections in the data allows fitting only models with few predictors.

We confirmed the large effect sizes of the C282Y and H63D SNPs and their interaction. Similarly, we could again see the significant risk increasing effect of age (except for the Best FinnGen logistic model), and that females are less likely to develop hemochromatosis. But on the other hand, we found a risk increasing interaction between being the postmenopausal female and the C282Y variables. This is especially visible in the meta-analysed models for the significant interaction between females and C282Y homozygotes.

In addition, we found that the SNP Causal1 had clear risk effect in all FinnGen models, whose effect is independent of C282Y, as shown in the logistic regression of hemochromatosis in data where C282Y-carriers are removed, see Figure S10. However, we were not able to replicate this finding due to this SNP being too rare in UKBB.

The results of a sensitivity analysis where the hemochromatosis cases were required to also have at least one venesection procedure performed are shown in Figures S11 and S12.

### Personal level risk estimates

We modeled the individual-level hemochromatosis risk as the probability of hemochromatosis diagnosis during follow-up given by a logistic regression. To evaluate the utility of the prediction models, we computed the hemochromatosis risks for several interesting predictor combinations for the Comparison and Best FinnGen logistic models.

In Figure S14 and in Table S10 the large risk increasing effect of C282Y in the Comparison model can be seen. Also, the effect of sex is large.

In the risk table of the best FinnGen model in Figure 4 and in Table S11 the large protective effect of blood donation and the risk effect of C282Y and Causal1 SNPs were seen. Also, the premenopausal females are at much lower risk of hemochromatosis than postmenopausal females.

**Figure 4.**
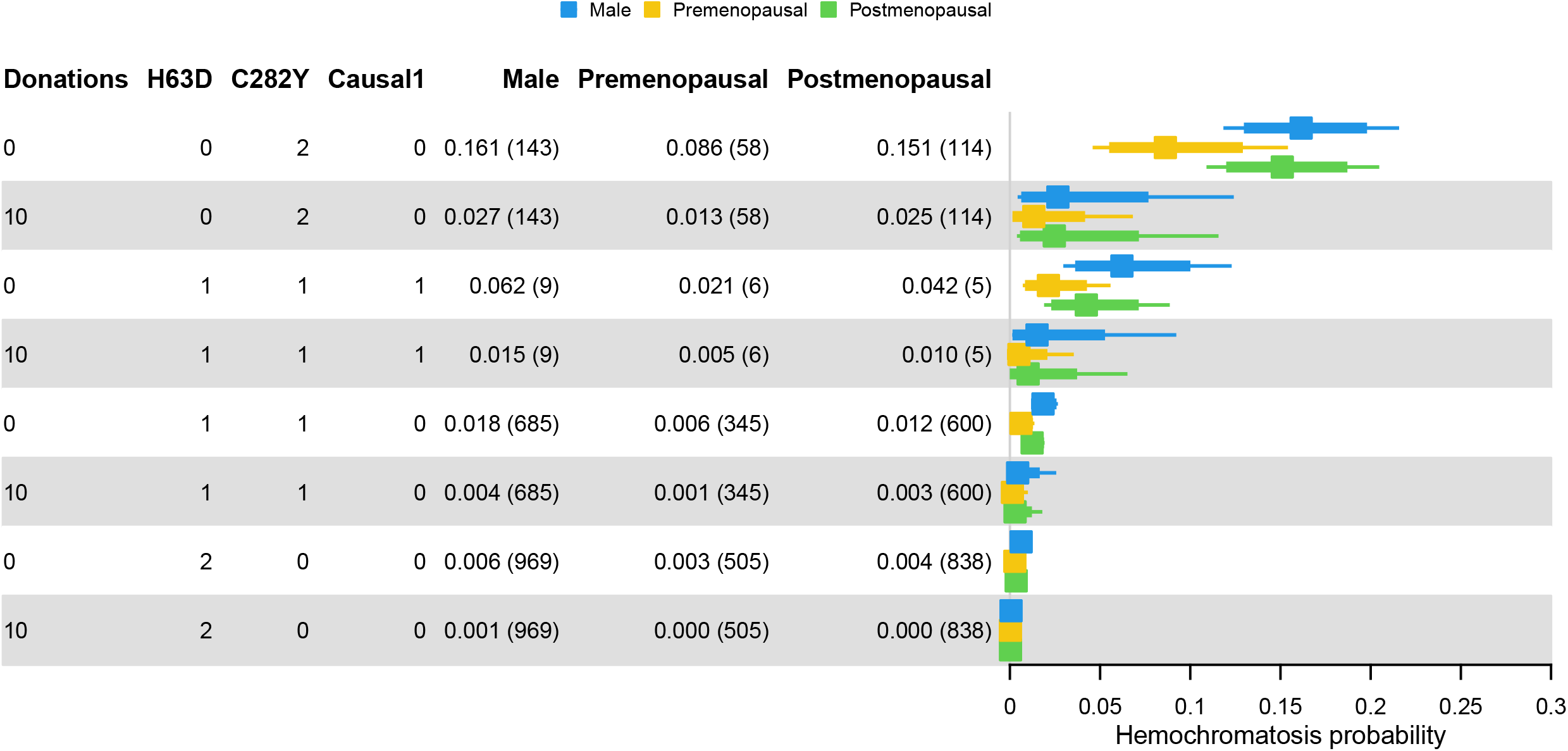
Risk table of the best FinnGen incidence model. The hemochromatosis risk (x axis) with both 80% and 95% confidence intervals for different predictor combinations. Predictor variables not shown were held fixed at mode or mean. The predictor combinations are ordered by the maximum risk over sexes. In parentheses the counts of genetic predictor combinations in full data are shown (predictors not shown were ignored when counting).

For C282Y homozygotes without the Causal1 variant the hemochromatosis risk drops statistically significantly. For non-donors the estimates are 0.161 (80% CI 0.133–0.195), 0.086 (0.058–0.126), and 0.151 (0.123–0.184), for males, premenopausal, and postmenopausal females, respectively. For individuals who donate 10 times in five years the respective estimates are 0.027 (0.009–0.074), 0.013 (0.004–0.039), and 0.025 (0.009–0.068), for males, premenopausal, and postmenopausal females, respectively.

Moreover, for C282Y homozygotes who donate the maximum number of donations in five years (20 for males and 15 for females), hemochromatosis risks are 0.004 (0.000–0.090), 0.005 (0.000–0.055), and 0.009 (0.001–0.098), for males, premenopausal and postmenopausal females, respectively. That is, the risks get smaller, but the confidence intervals get wider.

## Discussion

The subpopulations of blood donors and non-blood donors in FinnGen seem to be quite different. For example, the hemochromatosis prevalence was 0.12% in non-donors and 0.07% in donors (2-sample test for equality of proportions with continuity correction, p-value 0.00359, 95% CI for the difference [-0.07%, -0.02%]). There is also a marked difference in C282Y prevalence in hemochromatosis cases with MAF being 39.9% in non-donors and 64.1% in donors (p-value 4.024×10^-5^, 95% CI for the difference [-36.4%, -12.8%]). This could be due to an individual diagnosed with hemochromatosis informing his relatives of the risk, causing the relatives to start donating blood as a preventive measure. This hypothesis agrees with the detected enrichment of first-degree relatives among blood donors (15).

Even though, for example, Pilling et al have fitted separate models for males and females, in our data the onset and prevalence of hemochromatosis did not show large differences between males and females (Welch two-sample t-test for the difference of mean diagnosis age, male mean 56, female mean 55.2, p-value 0.514). Hence, we decided to fit a single model that includes sex as predictor, as this maximizes the statistical power of the fitted models in our setting of small number of hemochromatosis cases. Also, as the minimum recruitment age in the UKBB was 40, it was not sensible to split females into pre and postmenopausal in the UKBB data nor in the Comparison models.

The 67 SNPs putatively interacting with C282Y discovered by Regenie did not improve the prediction performance significantly, see Figure S6. Hence, we did not further investigate the contribution or mechanism of individual SNPs.

There is a large difference in minor allele frequencies between cases and controls in both C282Y and H1-2 in both FinnGen and UKBB. This indicates that they might be usable in predicting the hemochromatosis diagnosis. Including both SNPs in the logistic models caused problems with collinearity, the LD between H1-2 and H63D being 0.23 R^2^, see Figure S3. Hence, we excluded H1-2 from the logistic models, since it made interpretation of the effect sizes problematic. However, the SNP H1-2 was included in the All interactions model where it did not show large effect, see Figure S9 and Table S8 of variable importances, and in the non-C282Y-carrier model, where the effect of H1-2 was insignificant, see Figure S10.

The minor allele frequency is greater in UKBB for both C282Y (FinnGen 3.7%, UKBB 7.4%) and H63D (FinnGen 11%, UKBB 15.1%). However, in hemochromatosis cases the MAF of H63D is higher in FinnGen. This is probably a consequence of the fact that the frequency of C282Y homozygotes is much higher in UKBB, see panels D and E of Figure 2. As C282Y and H63D variants cannot exist in the same chromosome, this causes lower MAF of H63D in UKBB.

The low percentage of *HFE*-mutation carriers among hemochromatosis cases (75%) could be due to the diagnosis being secondary hemochromatosis due to some other disease, or due to a non-*HFE* form of hemochromatosis, with mutations in *HJV, HAMP, TRF2* or *SLC40A1* genes.

The Causal1 SNP is in gene *CASC15* (cancer susceptibility 15), which has been demonstrated to be associated with hepatocellular cancer (16). Our analysis in the non-C282Y-carriers showed its effect on hemochromatosis independently from the C282Y (see Figure S10) and it is not in linkage with C282Y (see Figure S3). Unfortunately, it is quite rare a variant: all the three detected causal SNPs had only 0.3% carriers, in FinnGen, and in the UKBB they were even more rare, which caused us to reject these SNPs from the comparison models.

The SNP HLA3, located in the HLA area in gene *HLA-DQA1*, has been shown by Clancy et al to be strongly associated with being a blood donor (14). Neither the variant itself nor any of the alleles of its containing gene *HLA-DQA1* (see Figure S4) was associated with hemochromatosis, but there was a slight protective interaction between HLA3 and C282Y in FinnGen and in UKBB just below the significance threshold.

The Figure 4 suggests that at least 10 blood donations per five years is enough to reduce the hemochromatosis risk when carrying C282Y in homozygous form to the level of carrying it as heterozygous. This is doable since in Finland males can donate 4 times a year and women 3 times a year. An exception to this rule are women between ages 18 and 25, who are recommended to donate only once per year, but on the other hand, young women are not at risk of hemochromatosis.

As the penetrance of C282Y SNP is roughly 20% in FinnGen, there must be some other factors affecting the penetrance. There has been discussion about interacting variants that modify the effect of C282Y, see for instance (5). However, we did not find them to have a large effect. The environmental factors are important as well: alcohol and red meat consumption have been shown to affect hemochromatosis (3), but unfortunately, this information is not available in FinnGen.

The prevalence of hemochromatosis in our modeling data was 0.1% in FinnGen and 0.4% in UKBB, while the MAFs of C282Y was 3.7% in FinnGen and 7.4% in UKBB. The disparity between the ratio of prevalences and the ratio of MAFs could indicate that hemochromatosis is underdiagnosed in Finland. This is further supported by the fact that the five preceding diagnoses in our models significantly predict hemochromatosis.

Pilling et al propose that large proportions of C282Y homozygotes eventually develop a hemochromatosis. They base this claim on the survival curves in C282Y homozygotes shown by Gallego et al, which show that by age of 90, the penetrance was nearly 50% among males and 25% among females. As their data was based on only 98 homozygotes with separate survival curves for males and females without confidence bands or numbers at risk, the estimates can be questioned. Similar analysis in FinnGen, stratified by sex and C282Y dosage, could not show a significant difference in survival from hemochromatosis diagnosis between male and female homozygotes, see Figure S13. The survival at age of 90 was 66% for males (5 at risk, 95% CI 58%–75%) and 66% (7 at risk, 95% CI 59%–74%).

The importance of blood donation in hemochromatosis risk shown in our analyses underlines the importance taking special attention to our blood donors diagnosed with hemochromatosis or at risk of developing the disease. Given that genetic data is coming increasingly available, our results highlight the need to make C282Y homozygotes aware of their risk and possibility of maintaining their health and helping others by the habit of blood donation. Previously at least eight blood establishments have allowed shorter donation intervals for *HFE* variant carriers or hemochromatosis patients (17). Also, making the therapeutic donors identify themselves more as donors than patients can have benefits for the blood establishment and the donor, such as increased commitment to donation and spreading good word about blood donation (18). In addition, the iron supplementation, which is automatically offered for frequent donors by the blood service in Finland, should not be given to C282Y homozygotes.

In conclusion, even though we could not find strong effects among the set of discovered novel modifiers of the C282Y variant, we did find a novel variant, Causal1, with risk effect independent from the C282Y variant. With the additional quantification of the strong effect of blood donation,our aims of research were largely successful. Due to the Causal1 variant being rare in UKBB and lack of blood donation information, these findings could not be replicated. But as the effects of other variants were remarkably similar in FinnGen and in UKBB, which represent genetically quite dissimilar populations, the results should be broadly applicable. And when the biology related to the HFE variants should work the same way, the effect of blood donation should also be generalizable.

## Supporting information

List of FinnGen members

## Data availability statement

Data will be made available on request.

## Code availability

All analysis R code will be made available at GitHub: https://github.com/FRCBS/hemochromatosis_modeling_article

## Acknowledgments

All participants of the FinnGen research project are listed in the supplemental table (FinnGen banner).

## Author contributions

JT, JC, JR and MA designed the study; FinnGen performed the hemochromatosis GWAS and finemapping while JT performed all other analyses; FÅ provided medical expertise; JT wrote the initial version of the manuscript. All authors read and critically commented the manuscript.

## Ethical approval

Ethical approval for this study (Ethical Committee statement number for the FinnGen study is Nr HUS/990/2017) was provided by the Coordinating Ethics Committee of the Hospital District of Helsinki and Uusimaa (HUS), and informed consent was obtained from all study subjects, for details see the supplemental methods.

