## Supplemental text and figures for "Quantifying risk modifiers of hereditary hemochromatosis using genomic and electronic health record data from FinnGen and UK Biobank"

2025-09-23

### Table of contents

**Ethics statement and materials & methods** S2

**Methods** S3

**Results** S4

**Acknowledgements** S5

**Figures** S5

**References** S20

### List of Figures

S1 Manhattan plot of the hemochromatosis GWAS in FinnGen . . . . . . . . . . . S6
S2 QQ plot of the hemochromatosis GWAS and the genomic inflation factor lambda. S7
S3 The linkage disequilibrium among SNPs and significant HLA alleles. . . . . . . S8
S4 Assocation analysis between hemochromatosis and HLA alleles . . . . . . . . . S9
S5 Results of the C282Y-interaction hemochromatosis GWASes . . . . . . . . . . . S10
S6 Prediction performances of the models . . . . . . . . . . . . . . . . . . . . . . . . S11
S7 The performance curves for the FinnGen hemochromatosis incidence models . . S12

S8 The variable importances of the XGBoost version of the Best FinnGen model. . S13
S9 The variable importances of the XGBoost version of the All interactions model S14
S10 Best FinnGen model with C282Y-carriers removed . . . . . . . . . . . . . . . . . S15
S11 Alternative version of the Comparison model . . . . . . . . . . . . . . . . . . . S16
S12 Alternative version of the Best FinnGen model . . . . . . . . . . . . . . . . . . S17
S13 Kaplan-Meier curves of time to hemochromatosis diagnosis stratified by sex and HFE genotype. . . . . . . . . . . . . . . . . . . . . . . . . . . . . . . . . . . S18
S14 Risk table of the FinnGen comparison incidence model . . . . . . . . . . . . . . . S19

### Ethics statement and materials & methods

Study subjects in FinnGen provided informed consent for biobank research, based on the Finnish Biobank Act. Alternatively, separate research cohorts, collected prior the Finnish Biobank Act came into effect (in September 2013) and start of FinnGen (August 2017), were collected based on study-specific consents and later transferred to the Finnish biobanks after approval by Fimea (Finnish Medicines Agency), the National Supervisory Authority for Welfare and Health. Recruitment protocols followed the biobank protocols approved by Fimea. The Coordinating Ethics Committee of the Hospital District of Helsinki and Uusimaa (HUS) statement number for the FinnGen study is Nr HUS/990/2017.

The FinnGen study is approved by Finnish Institute for Health and Welfare (permit numbers: THL/2031/6.02.00/2017, THL/1101/5.05.00/2017, THL/341/6.02.00/2018, THL/2222/6.02.00/2018, THL/283/6.02.00/2019, THL/1721/5.05.00/2019 and THL/1524/5.05.00/2020), Digital and population data service agency (permit numbers: VRK43431/2017-3, VRK/6909/2018-3, VRK/4415/2019-3), the Social Insurance Institution (permit numbers: KELA 58/522/2017, KELA 131/522/2018, KELA 70/522/2019, KELA 98/522/2019, KELA 134/522/2019, KELA 138/522/2019, KELA 2/522/2020, KELA 16/522/2020), Findata permit numbers THL/2364/14.02/2020, THL/4055/14.06.00/2020, THL/3433/14.06.00/2020, THL/4432/14.06/2020, THL/5189/14.06/2020, THL/5894/14.06.00/2020, THL/6619/14.06.00/2020, THL/209/14.06.00/2021, THL/688/14.06.00/2021, THL/1284/14.06.00/2021, THL/1965/14.06.00/2021, THL/5546/14.02.00/2020, THL/2658/14.06.00/2021, THL/4235/14.06.00/2021, Statistics Finland (permit numbers: TK-53-1041-17 and TK/143/07.03.00/2020 (earlier TK-53-90-20) TK/1735/07.03.00/2021, TK/3112/07.03.00/2021) and Finnish Registry for Kidney Diseases permission/extract from the meeting minutes on 4th July 2019.

The Biobank Access Decisions for FinnGen samples and data utilized in FinnGen Data Freeze 12 include: THL Biobank BB2017_55, BB2017_111, BB2018_19, BB_2018_34, BB_2018_67, BB2018_71, BB2019_7, BB2019_8, BB2019_26, BB2020_1, BB2021_65, Finnish Red Cross Blood Service Biobank 7.12.2017, Helsinki Biobank HUS/359/2017, HUS/248/2020, HUS/430/2021 §28, §29, HUS/150/2022 §12, §13, §14, §15, §16, §17, §18, §23, §58, §59, HUS/128/2023 §18, Auria Biobank AB17-5154 and amendment #1 (August 17 2020) and amendments BB_2021-0140, BB_2021-0156 (August 26 2021, Feb

2 2022), BB_2021-0169, BB_2021-0179, BB_2021-0161, AB20-5926 and amendment #1 (April 23 2020) and it´s modifications (Sep 22 2021), BB_2022-0262, BB_2022-0256, Biobank Borealis of Northern Finland_2017_1013, 2021_5010, 2021_5010 Amendment, 2021_5018, 2021_5018 Amendment, 2021_5015, 2021_5015 Amendment, 2021_5015 Amendment_2, 2021_5023, 2021_5023 Amendment, 2021_5023 Amendment_2, 2021_5017, 2021_5017 Amendment, 2022_6001, 2022_6001 Amendment, 2022_6006 Amendment, 2022_6006 Amendment, 2022_6006 Amendment_2, BB22-0067, 2022_0262, 2022_0262 Amendment, Biobank of Eastern Finland 1186/2018 and amendment 22§/2020, 53§/2021, 13§/2022, 14§/2022, 15§/2022, 27§/2022, 28§/2022, 29§/2022, 33§/2022, 35§/2022, 36§/2022, 37§/2022, 39§/2022, 7§/2023, 32§/2023, 33§/2023, 34§/2023, 35§/2023, 36§/2023, 37§/2023, 38§/2023, 39§/2023, 40§/2023, 41§/2023, Finnish Clinical Biobank Tampere MH0004 and amendments (21.02.2020 & 06.10.2020), BB2021-0140 8§/2021, 9§/2021, §9/2022, §10/2022, §12/2022, 13§/2022, §20/2022, §21/2022, §22/2022, §23/2022, 28§/2022, 29§/2022, 30§/2022, 31§/2022, 32§/2022, 38§/2022, 40§/2022, 42§/2022, 1§/2023, Central Finland Biobank 1-2017, BB_2021-0161, BB_2021-0169, BB_2021-0179, BB_2021-0170, BB_2022-0256, BB_2022-0262, BB22-0067, Decision allowing to continue data processing until 31st Aug 2024 for projects: BB_2021-0179, BB22-0067,BB_2022-0262, BB_2021-0170, BB_2021-0164, BB_2021-0161, and BB_2021-0169, and Terveystalo Biobank STB 2018001 and amendment 25th Aug 2020, Finnish Hematological Registry and Clinical Biobank decision 18th June 2021, Arctic biobank P0844: ARC_2021_1001.

### Methods

We use bgenix (version 1.1.7), qctool (2.2.0), tabix (1.16) and bcftools (1.18) to process genotype data. The additive hemochromatosis GWAS is done using Regenie version 2.2.4. Fine-mapping of causal variants is done using Susie version 0.11.92.2360607.

The HLA alleles are imputed using the previously developed population-specific models for classical [11] and non-classical [13] HLA genes based on HIBAG [17] versions 1.18.1 and 1.40.0, and haplotyping is performed using the gap R package version 1.5.3. Genome-wide search of putative variants that interact with C282Y is performed with Regenie version 3.2.3. We also experimented with MAPIT [3] to discover interactions on whole genome level but found it did not support binary responses in practice.

All further analysis is done in R version 4.4.2. The tidymodels framework [7] is used to fit the incidence models, survival models are fitted with the survival R package [14] version 3.7.0, and the venesection counts are fitted with the stats package version 4.4.2 (Poisson), MASS package [15] version 7.3.61 (negative binomial), and pscl package [16] version 1.5.9 (zero-inflated models).

The ciTools package [5] (version 0.6.1) is used to compute confidence intervals for the predicted probability of hemochromatosis, and the forestplot package [4] (version 3.1.6) is used to the visualize these confidence intervals. We use the shapviz package [9] (version 0.9.6) to estimate variable importances and analyze the interactions among the predictors of an XGBoost [2] incidence model using shap values [8]. A fixed-effect meta-analysis between the effect sizes of comparison models of FinnGen and UKBB is performed using the meta package [1] version 8.0.1. The Firth logistic regression is fitted using the logistf package [6] (version 1.26.1).

The data is split into train and test halves with approximately the same proportion of hemochromatosis cases, and unbiased performance measures are obtained by predicting on test data using models fitted on the train data. We use the area under the receiver operating characteristic (ROC) curve and the precision-recall (PR) curve as the performance measures based on which the best incidence and survival models are selected. After the model selection, the selected models are refitted on the full data to allow inference on the importance of the predictor variables.

The selection of the prior diseases was based on the survival analysis between the endpoints performed by FinnGen The method is described at Risteys, and repeated briefly here. All FinnGen endpoint pairs were considered in the analysis. The prior disease is considered as an exposure and the later disease as an outcome. The exposure is modeled as a time-dependent covariate. The follow-up time was from 1998-01-01 – 2021-12-31, or until diagnosis of the outcome endpoint or death. A case-cohort design was used for speed, with subcohort size of 10,000. The Cox regression was adjusted by sex and birth year. All previous diseases with statistically significant hazard ratio at level 0.05 with multiple test correction were selected as predictors.

In the sensitivity analysis, the resulting statistical problems due to low number of cases were handled using the Firth logistic regression.

### Results

We also performed association study of the hemochromatosis (264 cases, 210,007 controls) with the HLA genes (15 genes that had altogether 252 alleles) to understand the role of the HLA in the origin of hemochromatosis, which has been discussed in previous literature [10, 12], see supplemental Figure S4.A. Several associations were found, all in the genes of MHC class I, but after adjusting the GWAS with C282Y, all the associations disappeared, see Supplemental Figure S4.B.

As the extended HLA region was excluded from fine-mapping, we also selected all significant lead SNPs from the HLA area. However, we excluded one of them, since it was in too high LD with the other two ($R^2$ 0.6 and 0.66, see Figure S3).

The performance of the FinnGen hemochromatosis incidence models fitted on the train data are shown in Supplemental Figure S6 and Supplemental Table S5. The train, cross-validation and test performance measures agreed quite well with each other on models based on logistic regression, which indicated that no large overfitting had occurred. Whereas the XGBoost based models overfitted the train data considerably.

The baseline model seems to perform slightly better on the test data than the comparison model. This could be due to females being split into pre- and postmenopausal in the baseline model but not in the comparison model, since the menopausal status has important effect on the hemochromatosis incidence.

The flexibility of XGBoost model allows slight improvement over the logistic regression on the test data in the case of the Best FinnGen model, but it makes inference more difficult. Also, the All interactions model, which consists of the same variables as the Best FinnGen model complemented with the H1-2 variant and the 67 putatively C282Y-interacting variants, does not offer a large improvement over the simpler model. Hence, we concentrated on the logistic regression-based models for further analysis.

The ROC and PR curves corresponding to the performance analysis are shown in Supplemental Figure S7.

### Acknowledgements

We want to acknowledge the participants and investigators of the FinnGen study. The FinnGen project is funded by two grants from Business Finland (HUS 4685/31/2016 and UH 4386/31/2016) and the following industry partners: AbbVie Inc., AstraZeneca UK Ltd, Biogen MA Inc., Bristol Myers Squibb (and Celgene Corporation & Celgene International II Sàrl), Genentech Inc., Merck Sharp & Dohme LCC, Pfizer Inc., GlaxoSmithKline Intellectual Property Development Ltd., Sanofi US Services Inc., Maze Therapeutics Inc., Janssen Biotech Inc, Novartis AG, and Boehringer Ingelheim International GmbH. Following biobanks are acknowledged for delivering biobank samples to FinnGen: Auria Biobank, THL Biobank, Helsinki Biobank, Biobank Borealis of Northern Finland, Finnish Clinical Biobank Tampere, Biobank of Eastern Finland, Central Finland Biobank, Finnish Red Cross Blood Service Biobank, Terveystalo Biobank and Arctic Biobank. All Finnish Biobanks are members of BBMRI.fi infrastructure. Finnish Biobank Cooperative - FINBB is the coordinator of BBMRI-ERIC operations in Finland. The Finnish biobank data can be accessed through the Fingenious® services managed by FINBB.

### Figures

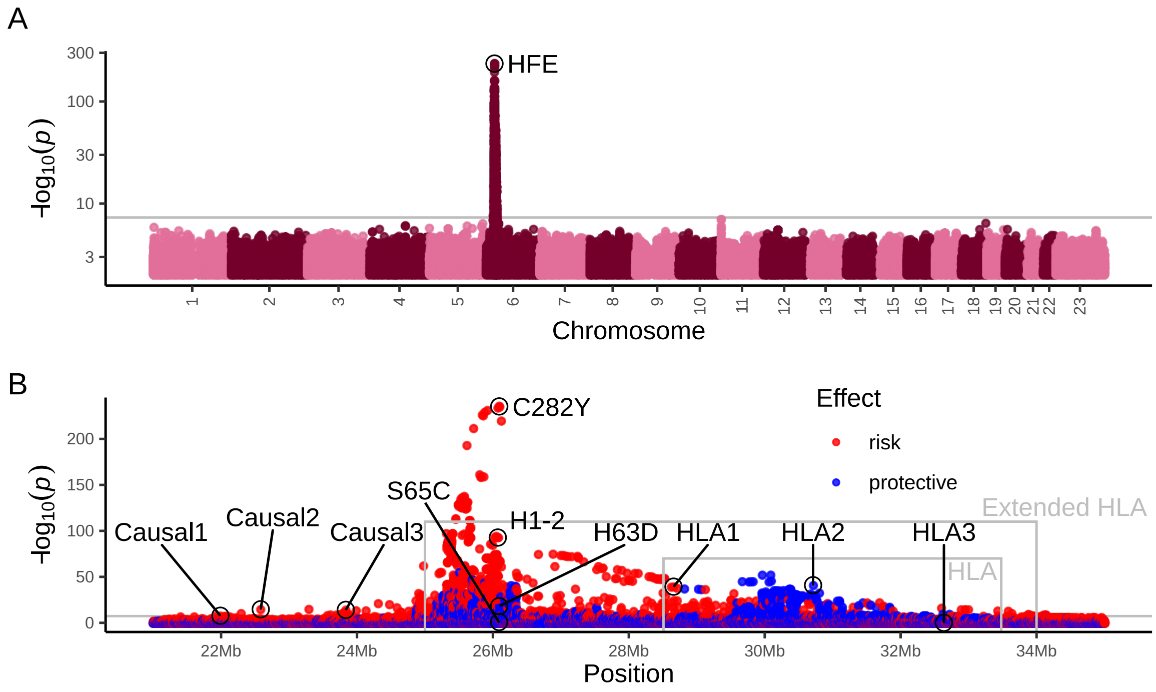


**Figure S1.** Manhattan plot of the hemochromatosis GWAS in FinnGen with 477 cases and 420,066 controls. A) Full Manhattan plot showing that the only peak resides near the HLA region in chromosome 6. B) Manhattan plot zoomed in to the peak region. The HLA area and the extended HLA area are shown as gray boxes. The genome-wide significance threshold of 5×10-8 is shown as gray horizontal line. The annotation in panel A shows the gene at the peak, and annotation in panel B shows the SNPs used as covariates in the statistical models.

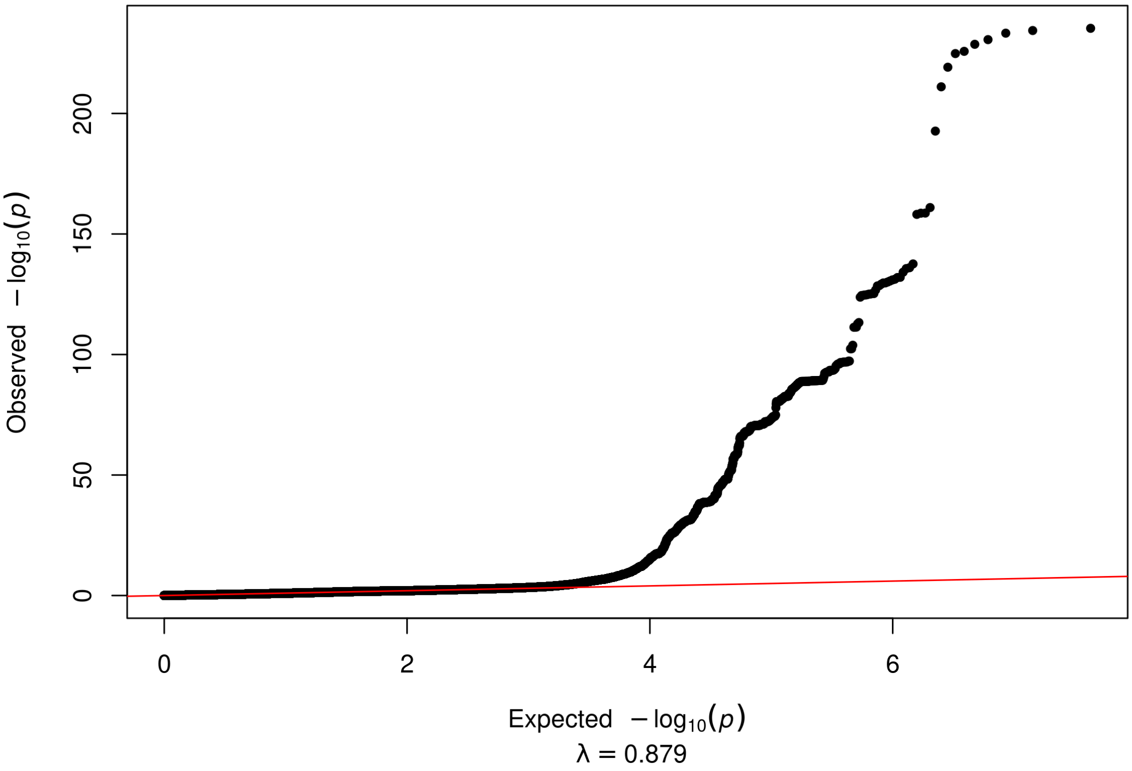


**Figure S2.** QQ plot of the hemochromatosis GWAS and the genomic inflation factor lambda.

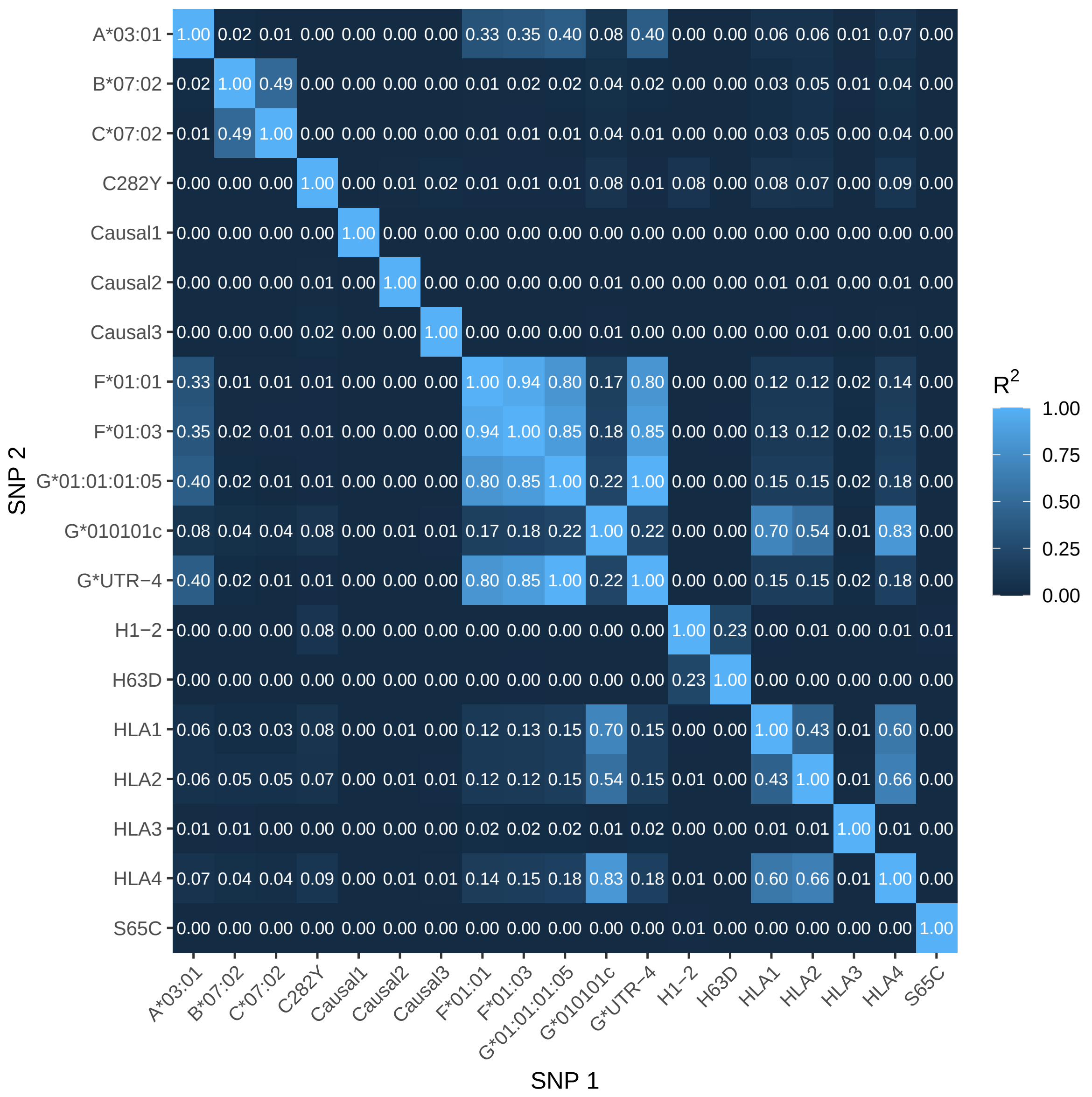


**Figure S3.** The linkage disequilibrium among SNPs and significant HLA alleles.

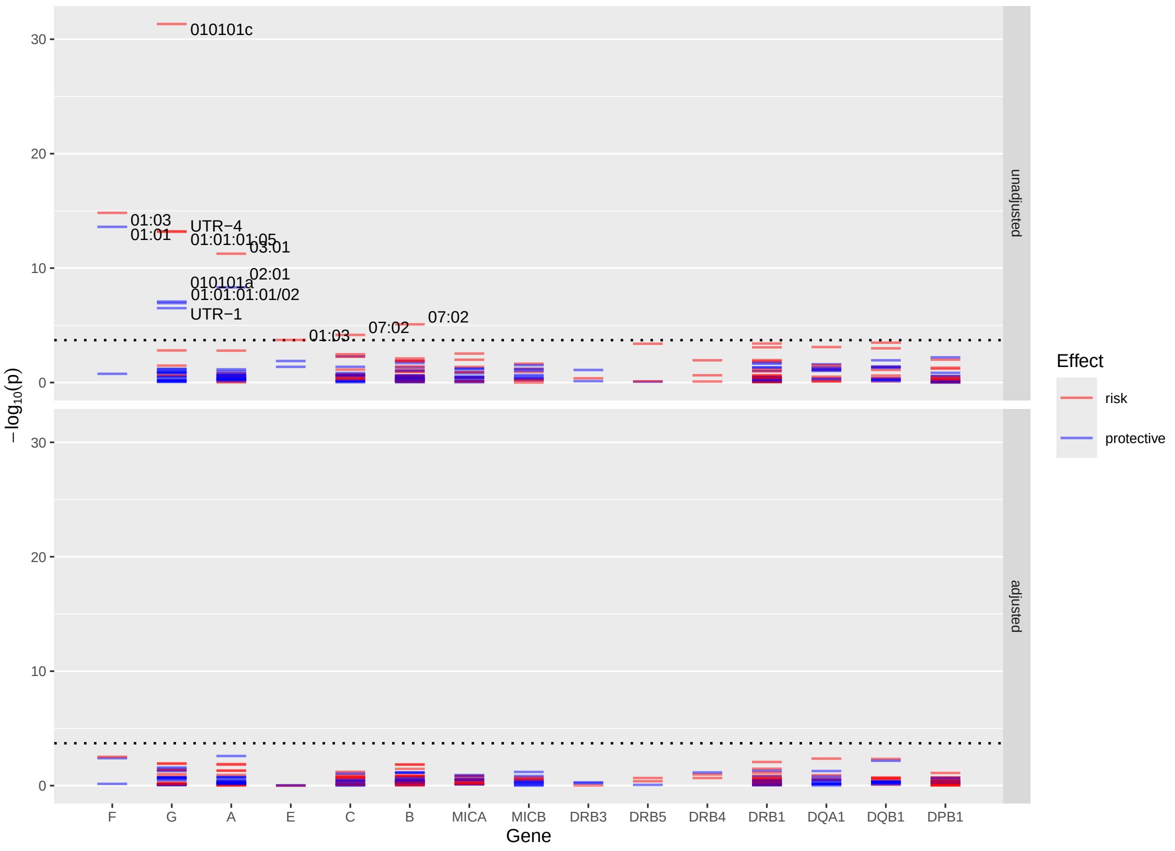


**Figure S4.** Results from association analysis of the hemochromatosis and HLA alleles. The analysis included fifteen HLA genes that had altogether 252 alleles. A) Analysis adjusted by sex, age, 10 PCs and genotyping batch. B) Analysis was additionally adjusted by the C282Y SNP. The multiple hypotheses corrected significance threshold is set to $-\log_{10}(0.05/252) \approx 3.7$ and marked with dotted lines in the panels.

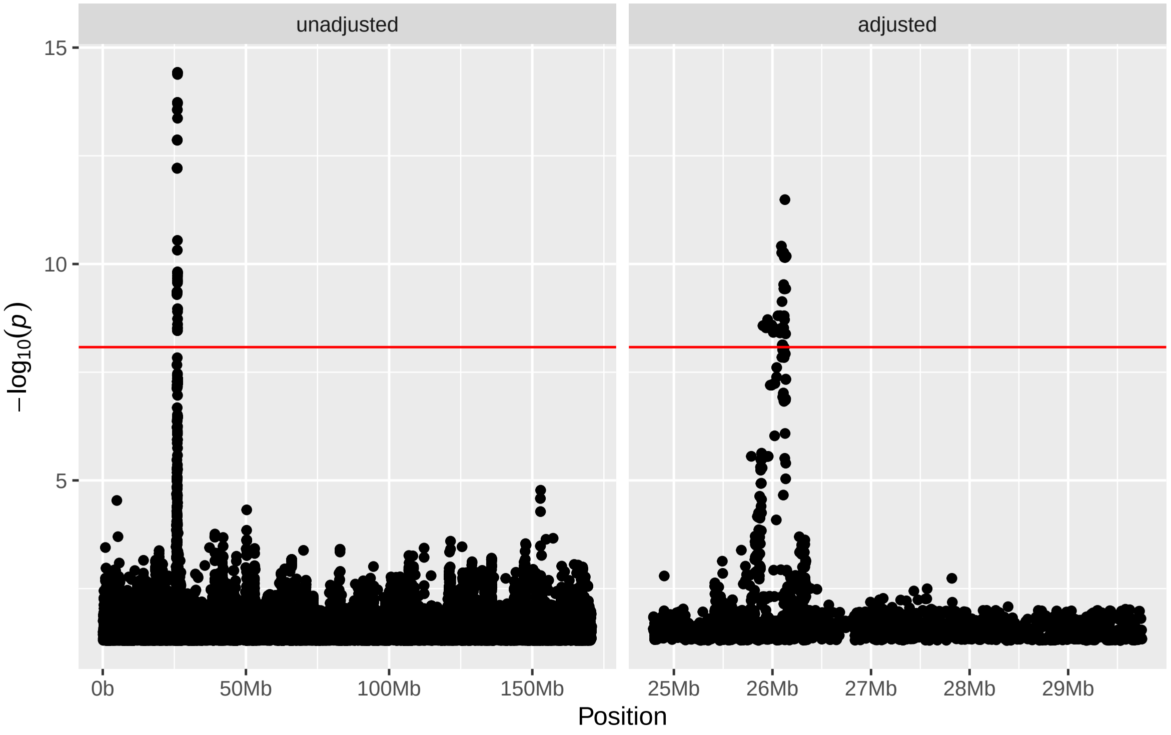


**Figure S5.** Results of the C282Y-interaction hemochromatosis GWASes (477 cases, 420,066 controls). On the left is the unadjusted GWAS, which found 42 significant interactions with C282Y. On the right is the GWAS adjusted by H63D, S65C, H1-2 and HLA3, which found 67 SNPSs that had significant interaction with C282Y.

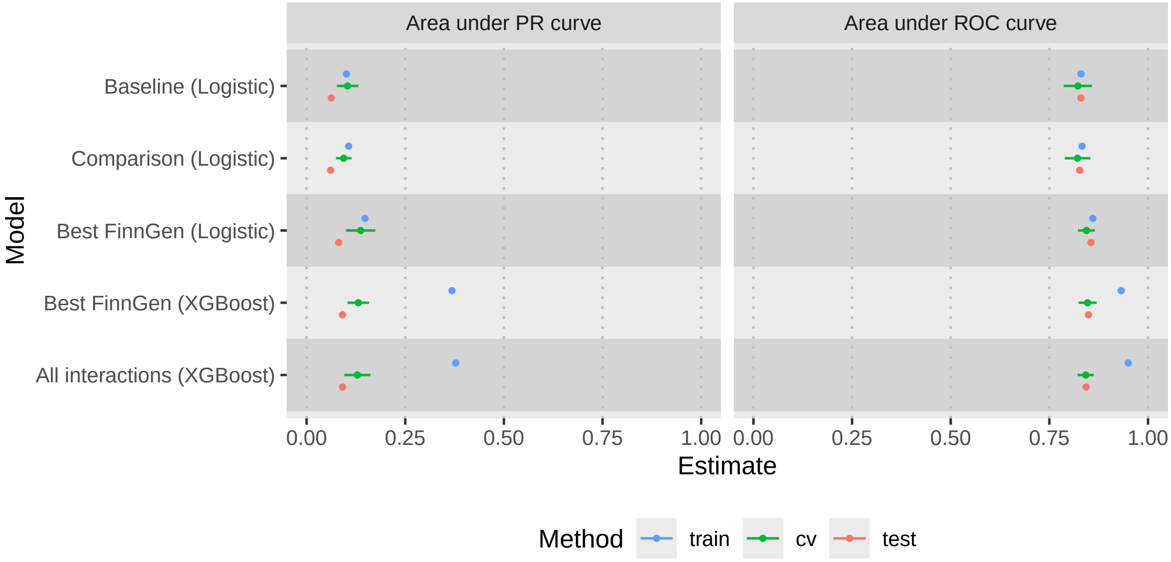


**Figure S6.** Prediction performances of the models of hemochromatosis incidence fit on the FinnGen train data (264 cases and 210,007 controls). The performances are evaluated on the train data, estimated using cross-validation, and on the held-out test data (213 cases and 210,059 controls). Ten-fold cross-validation (CV) was used to estimate the performance on new data. The error bars show the 95% confidence intervals for the cross-validation estimates.

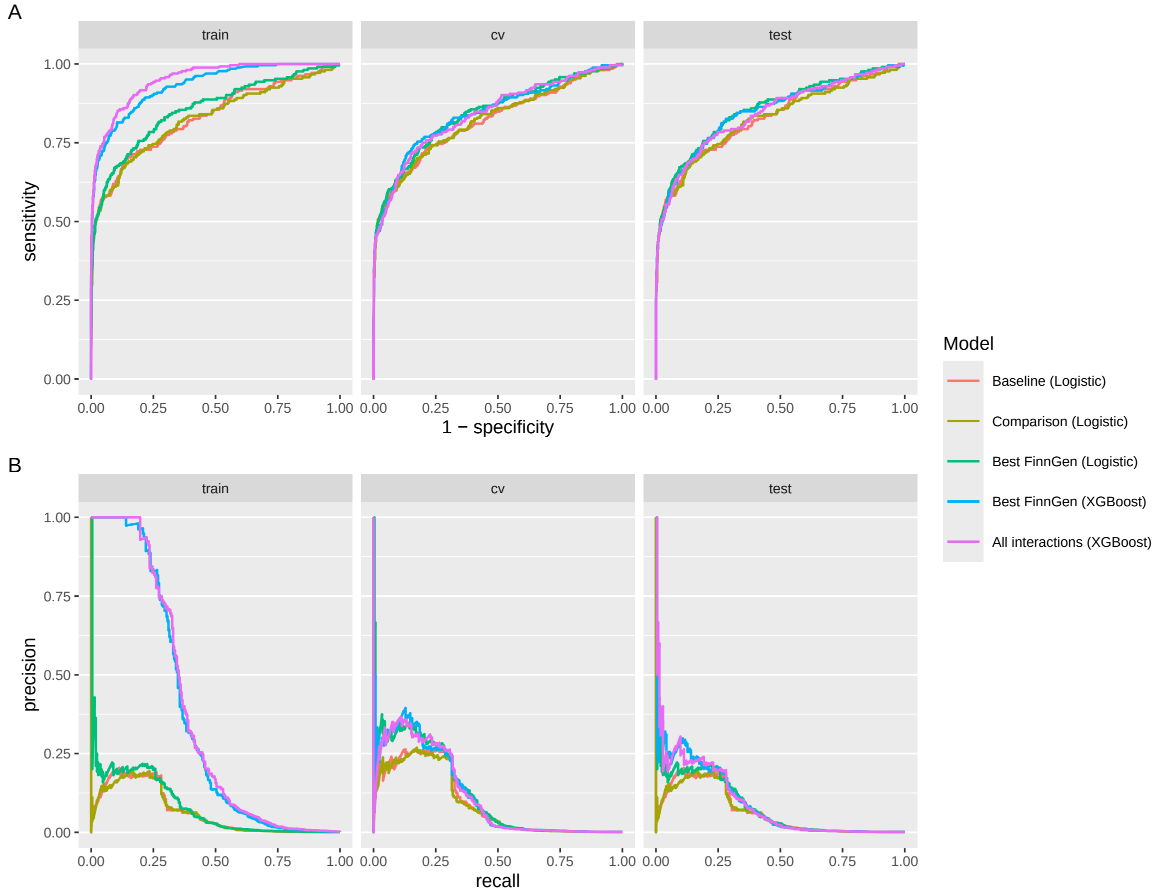


**Figure S7.** The performance curves for the FinnGen hemochromatosis incidence models. A) Receiver operating characteristic curves. B) Precision-recall curves.

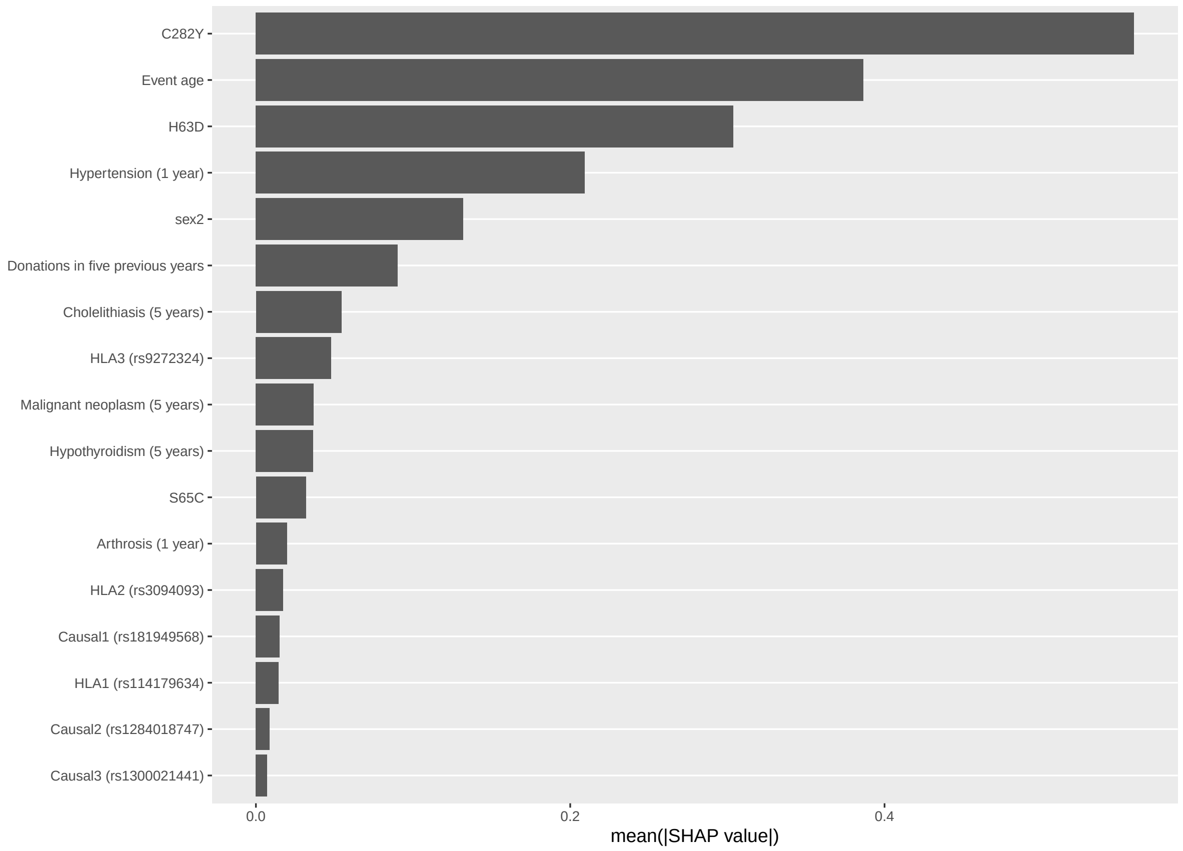


**Figure S8.** The variable importances of the XGBoost version of the Best FinnGen model.

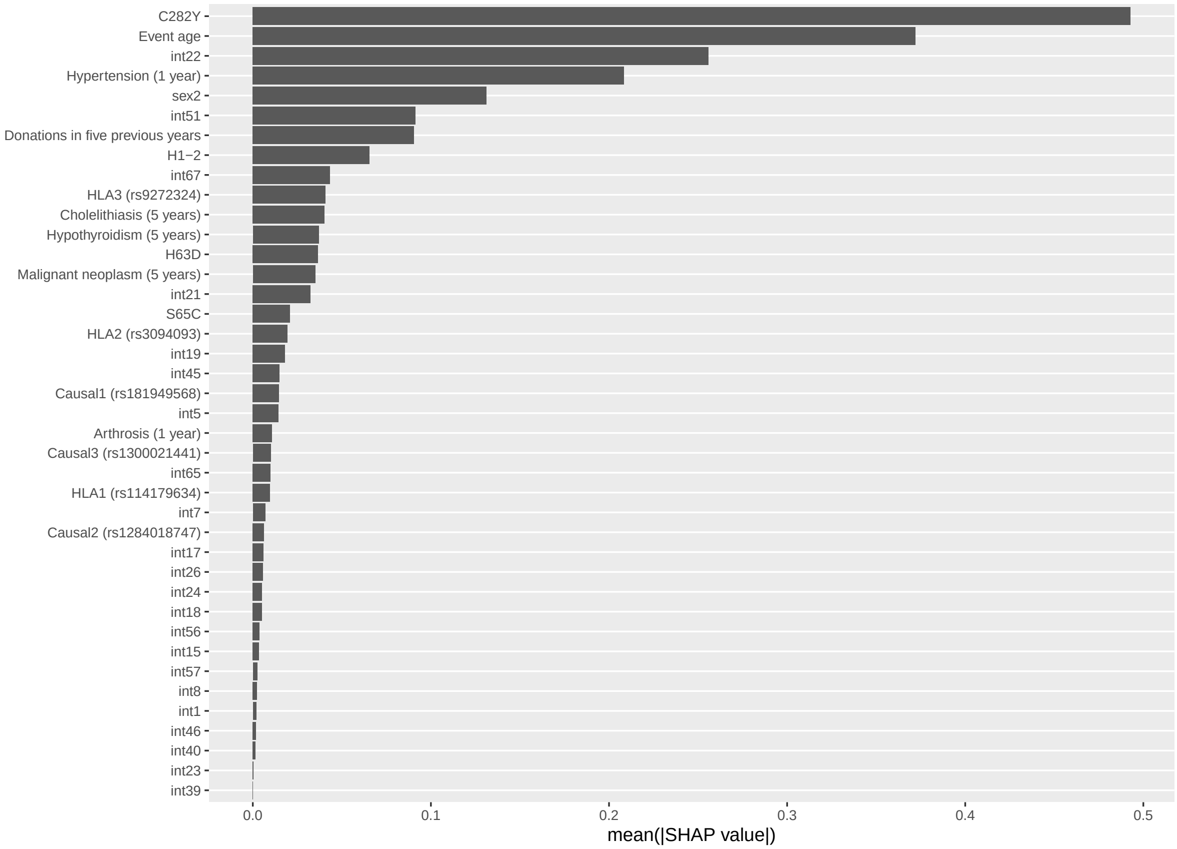


**Figure S9.** The variable importances of the XGBoost version of the All interactions model. Variables not shown had importance of zero.

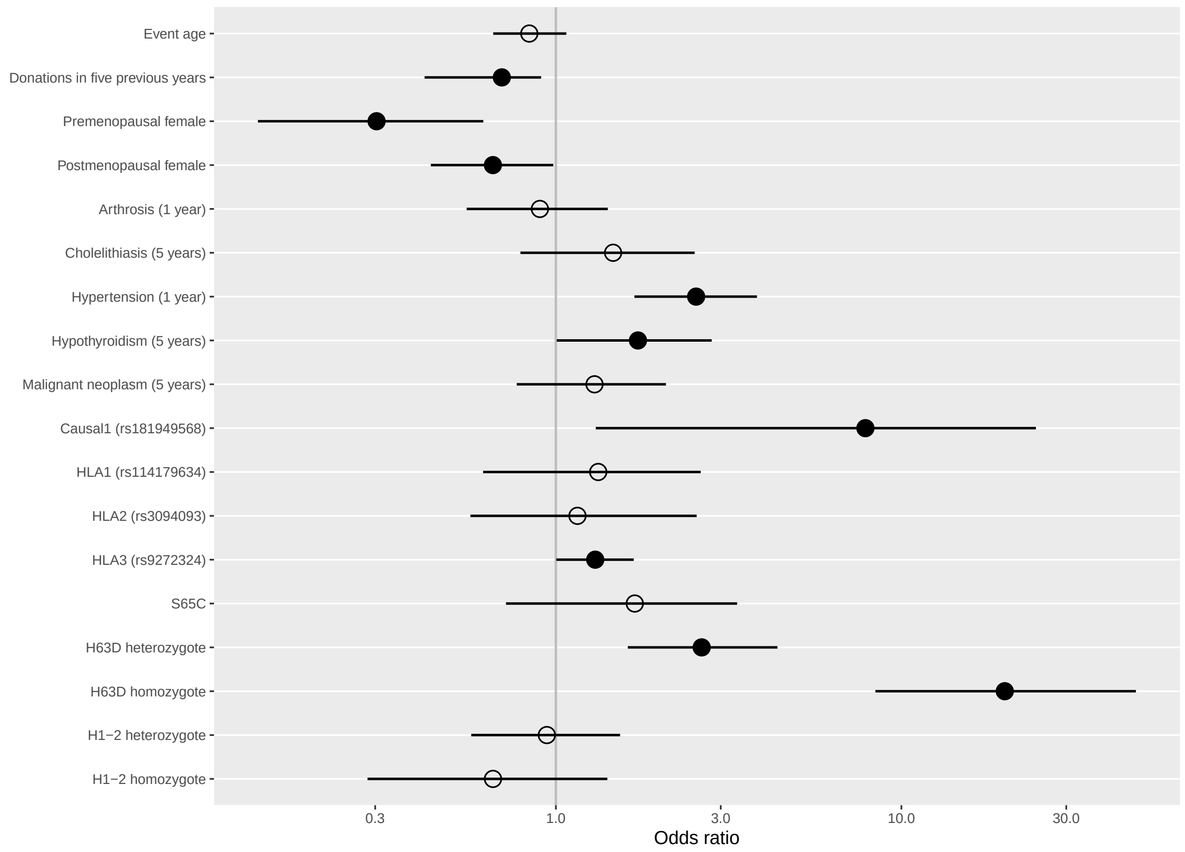


**Figure S10.** A version of the Best FinnGen model fitted on data (n=195,075, cases=120, prevalence=0.06%) where all C282Y carriers are removed.

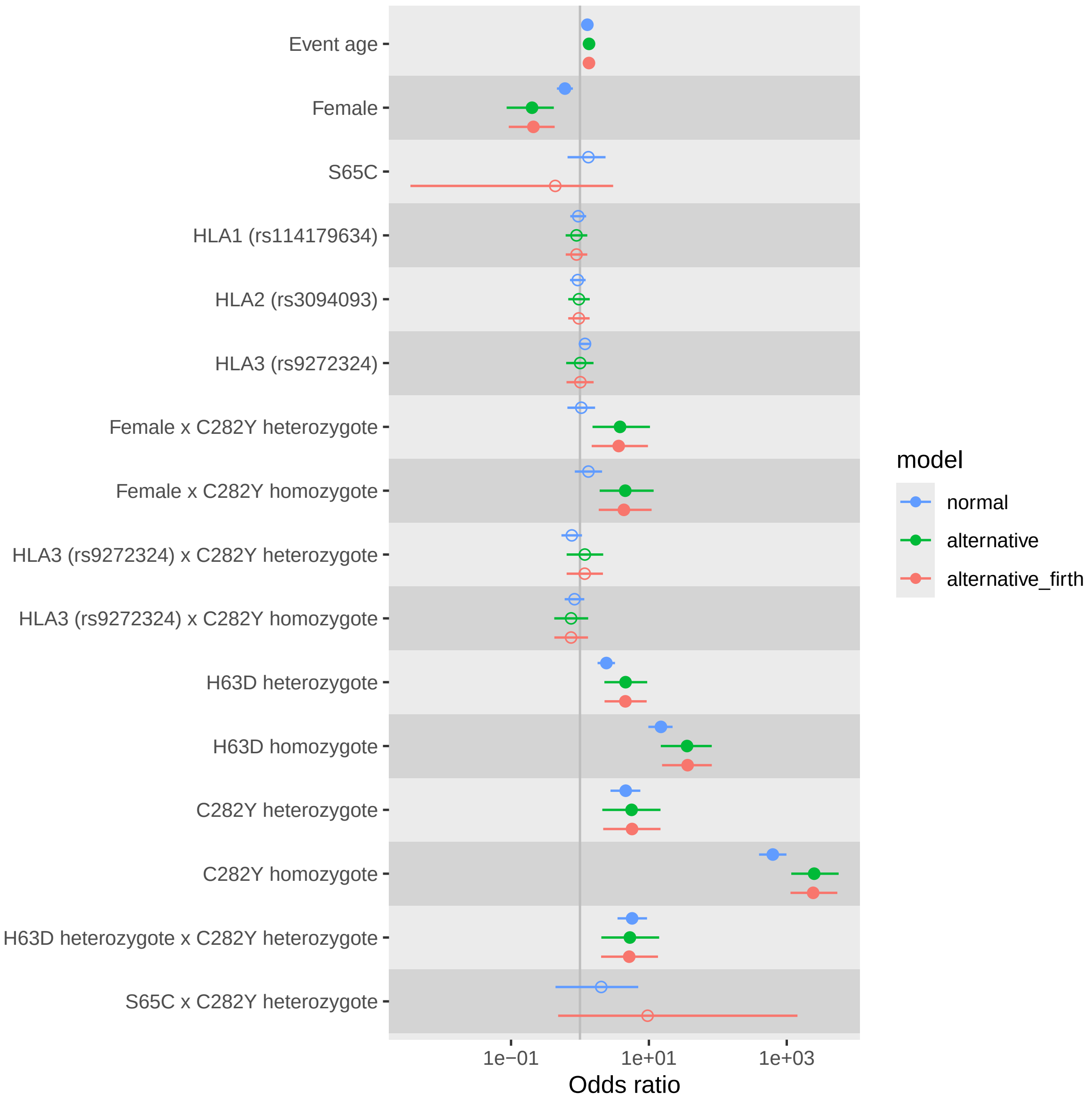


**Figure S11.** A version of the Comparison model fitted on data (n=420,251, cases=185, prevalence=0.04%) with alternative response definition where were cases were required to have at least one venesection performed. Individuals with a hemochromatosis diagnosis but no venesections were excluded from the data. The Firth logistic regression was also fitted for the alternative definition of the response to avoid estimation problems of coefficients of some variables due to low data. The odds ratio estimates and their 95% confidence intervals are shown for each variable. For comparison, the estimates are shown for the logistic regression of the normal hemochromatosis definition, the logistic regression of the alternative hemochromatosis definition, and the Firth logistic regression of the alternative hemochromatosis definition.

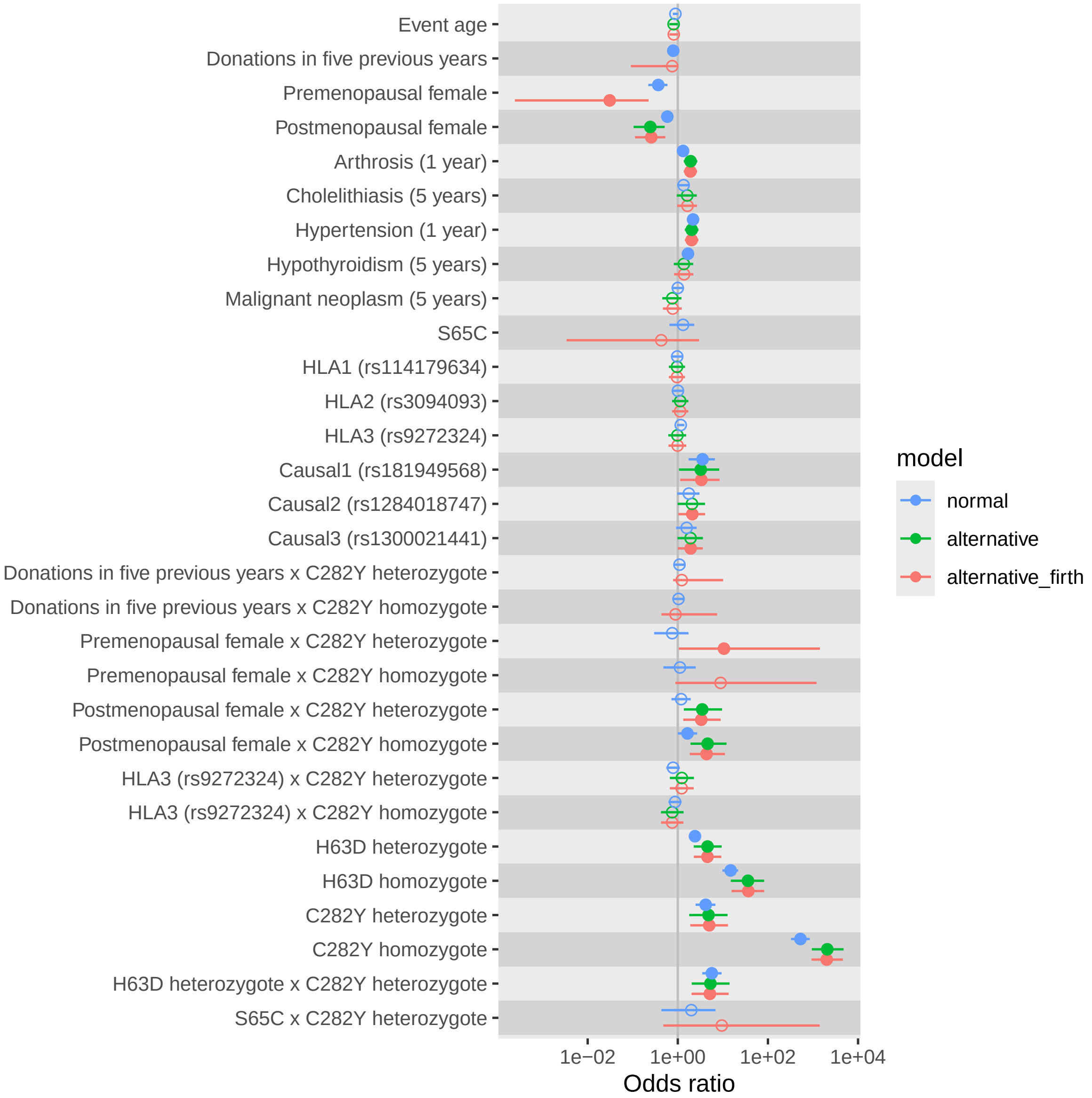


**Figure S12.** A version of the Best FinnGen model fitted on data (n=420,251, cases=185, prevalence=0.04%) where were cases are required to have at least one venesection performed. Individuals with a hemochromatosis diagnosis but no venesections were excluded from the data.

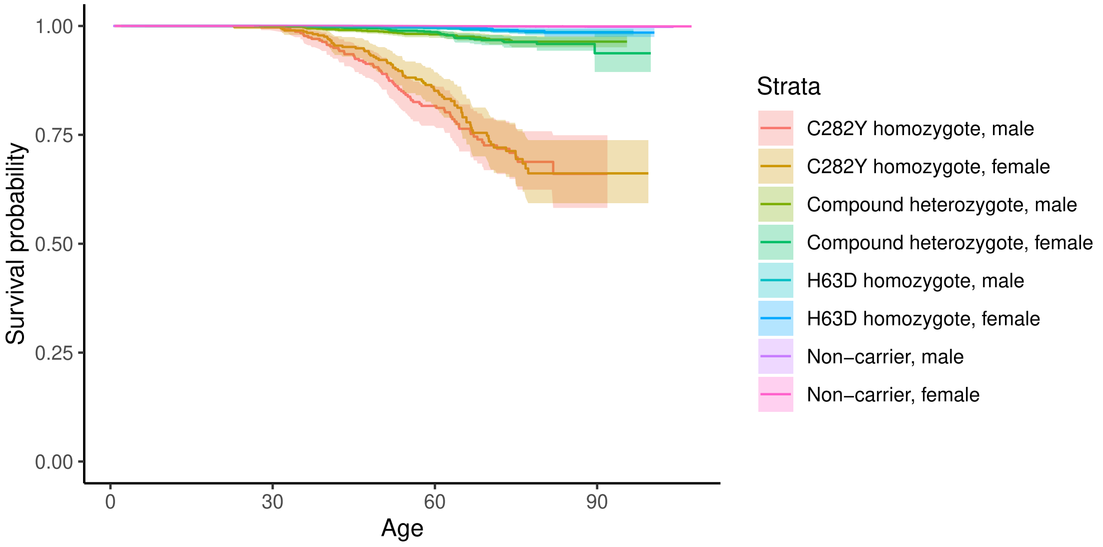


Number at risk

| Strata | 0 | 30 | 60 | 90 |
|---|---|---|---|---|
| C282Y homozygote, male | 314 | 304 | 172 | 5 |
| C282Y homozygote, female | 411 | 385 | 189 | 7 |
| Compound heterozygote, male | 1458 | 1370 | 863 | 26 |
| Compound heterozygote, female | 1979 | 1803 | 883 | 40 |
| H63D homozygote, male | 2223 | 2112 | 1405 | 39 |
| H63D homozygote, female | 3010 | 2755 | 1375 | 46 |
| Non−carrier, male | 173757 | 165194 | 104790 | 3187 |
| Non−carrier, female | 237391 | 217489 | 107173 | 3715 |

Age

**Figure S13.** Kaplan-Meier curves of time to hemochromatosis diagnosis stratified by sex and HFE genotype.

| H63D | C282Y | Male | Female |
|---|---|---|---|
| 0 | 2 | 0.215 (146) | 0.168 (180) |
| 1 | 1 | 0.025 (694) | 0.015 (956) |
| 2 | 0 | 0.008 (969) | 0.004 (1345) |
| 0 | 1 | 0.002 (5336) | 0.001 (7346) |
| 1 | 0 | 0.001 (15389) | 0.001 (20996) |
| 0 | 0 | 0.001 (60161) | 0.000 (82723) |

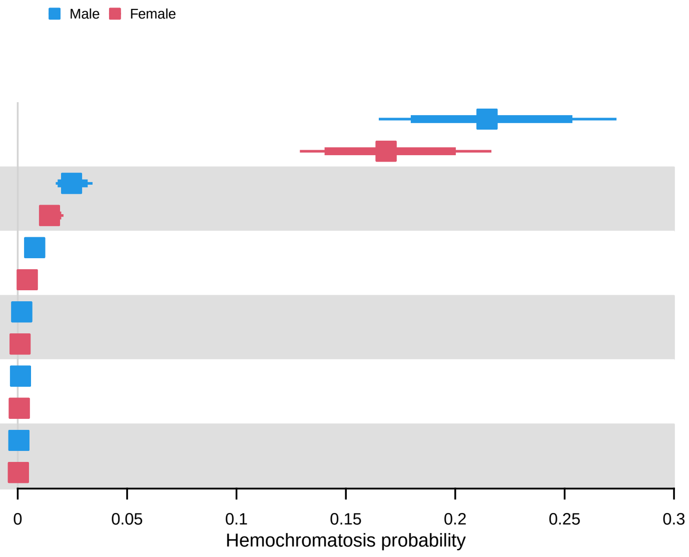


**Figure S14.** Risk table of the FinnGen comparison incidence model. The hemochromatosis risk (x axis) with both 80% and 95% confidence intervals for different predictor combinations. Predictor variables not shown were held fixed at mode or mean. The predictor combinations were ordered by the maximum risk over sexes. In parentheses the counts of predictor combinations in full data are shown (predictors not shown were ignored when counting).
